# Explanatory models of cancer and implications for healthcare-seeking: A qualitative study from an informal settlement in urban Dhaka, Bangladesh

**DOI:** 10.64898/2025.12.26.25342881

**Authors:** Md. Sajid Sultan Haque, Semonty Jahan, Lamisa Rahman, Nahitun Naher, Syed Masud Ahmed

## Abstract

Globally, over 19 million new cancer cases and 10 million deaths occurred in 2020, with 70% reported from low- and middle-income countries such as Bangladesh. Understanding community perceptions regarding cancer, including its Explanatory Model (EM), is essential for developing effective prevention and early detection strategies. This study explored the EM of cancer and its influence on healthcare-seeking behaviour among adults living in the Korail slum in Dhaka during October 2023 - January 2024. Fourteen in-depth interviews and two focus group discussions were conducted with a total of 26 participants (13 women and 13 men) aged 18–66 years, selected purposively to ensure diversity in age, gender, and occupation. Data were collected using semi-structured guides on perceptions and healthcare-seeking behavior related to cancer. Interviews were transcribed, coded, and analyzed thematically using inductive and deductive approaches guided by Kleinman’s Explanatory Model framework.

Three major themes emerged. First, cancer was perceived as fatal, incurable, and often divinely predetermined. Participants attributed it to fate, moral wrongdoing, food adulteration, pollution, and unhealthy habits such as smoking or betel nut use. Second, gendered and moralized beliefs shaped understandings of cancer causation, where women’s reproductive functions and menstrual or sexual practices were viewed as risks, while men’s substance use was associated with masculinity. These perceptions reinforced gendered blame and stigma. Third, although participants were aware of nearby cancer facilities, care-seeking was often delayed due to misconceptions, fatalistic beliefs, distrust in healthcare providers, financial hardship, and lack of awareness or health education.

Community EMs strongly influence cancer-related perceptions and health-seeking in urban poor settings. Customized, culturally grounded awareness and screening initiatives are essential to dispel misconceptions and improve early diagnosis. The findings offer critical insights for designing community-based cancer prevention strategies in similar low-income urban contexts.

## Introduction

Among the non-communicable diseases, cancer is one of the major global public health challenges [1]. Cancer is a fatal disease resulting from uncontrolled cell proliferation, which can be inherited or caused by external influences [2]. In 2020, an anticipated 19.3 million new cases of cancer and over 10.0 million cancer-related deaths occurred globally [3]. Approximately, 70% of these cancer-related deaths take place in low or middle income countries. It has been assumed that, by 2030, the incidence could reach 20 million [4]. WHO South-East Asian region is reported to have 2.37 million new cases and 1.53 million cancer-related deaths in 2022 [5].

Cancer is more than just a physical illness—it significantly impacts every aspect of a person’s life. Itis closely associated with emotional and psychological challenges such as depression, anxiety, and other serious mental health disorders. Cancer treatment is also financially challenging [6,7]. It does imply that the majority of cancers can be avoided through primary prevention, which involves altering environmental and behavioral factors. Additionally, secondary prevention, which involves building awareness, and early detection and intervention, is crucial for reducing cancer-related fatalities [8,9]. Studies have found a strong association between cancer-related knowledge and early reporting and detection, ultimately improving prognosis [10]. So, the importance of early detection and manageent cannot be overemphazized. Socio-cultural backgrounds and individual experiences significantly influence attitudes toward cancer and healthcare-seeking practices [11]. Global studies reveal diverse cultural interpretations of cancer [12, 13], often associating it with fear, magic, misfortune, or divine retribution, particularly in developing nations [14,15]. Some link illness to a perceived misalignment with God’s will, thereby undermining faith in healthy behaviors [14]. Over time, a shift towards hope and the belief in cancer survivorship has replaced older perceptions of cancer as a terminal and miserable condition. Often, these beliefs have been found to influence a person’s healthcare-seeking practice in a significant way [12]. Women may delay seeking medical attention for breast abnormalities due to various reasons, including fatalism, trust in divine healing, reluctance to discuss their condition with partners, or misconceptions about the condition [16–18]. Similarly, men may experience fear, humiliation, and stigma regarding prostate cancer early detection, hindering communication with healthcare experts, particularly when it affects physiological and reproductive health [19]. Because perceptions and beliefs about cancer vary widely, it is important to consider an individual’s perceptions and attitudes towards it, which may help us better understand their healthcare-seeking behaviours. To understand an individual’s lay health beliefs and behaviors regarding cancer, the “EM of illness”, developed by Arthur Kleinman and colleagues, is helpful [20]. This model is a concept that makes it easier for medical professionals and other service providers to comprehend the viewpoints and comprehension of patients regarding health-related matters [21].

In Bangladesh, cancer cases are also rising alarmingly. Approximately, 200,000 people receive a new cancer diagnosis each year [22]. It has been reported that cancer causes 12% of all deaths in the country [23]. The underlying factors for this rise of cancer incidence are due to factors like longer life expectancy, greater exposure to cigarette smoke and other air pollutants, a greater intake of calorie-dense, saturated fat meals, and less physical activity [24]. The most common cancers in Bangladesh are Breast, Lung, Cervical, Esophageal, lip, and oral cavity cancers [25]. The perceptions and attitudes towards cancer in the urban slums of Dhaka city is an area yet to explore. The majority of the scant cancer study data from Bangladesh currently available are quantitative and facility-based; very little focus has been given to the urban slum population to develop an in-depth understanding of their perspective on thedisease. The existing studies can provide us with a general idea of the disease burden but are insufficient to explore understanding and perceptions in the light of the EM of illness, which ultimately informs healthcare-seeking practices. in the informal urban setlements, e.g., in Dhaka City, access to healthcare and information is often limited. The “emic” perspective on cancer in the impoverished community will aid in collecting in-person data on the attitudes and viewpoints of the population, as well as traditional beliefs, health-seeking behaviours, etc. Thus, this study which was part of the requisite of Summative Learning Project of my Masters of Public Health degree, aimed to explore the community’s EM of illness regarding cancer and to understand how this may inform policymakers and practitioners in developing a culture-sensitive cancer prevention and detection model that affects the community’s efforts in early cancer detection and prevention through health-seeking practices.

## Methods

### Ethics Statement

Ethical approval for this study was obtained from the Institutional Review Board of BRAC JPGSPH (IRB No: MPH-2023-001). Participant recruitment and data collection were conducted between 22–28 November 2023. All participants (both men and women) were adults aged 18 years and above. Prior to participation, written informed consent was obtained from all participants. The consent process included an explanation of the study objectives, voluntary participation, confidentiality, photography and audio recording permission, using data for report and publication where anonymity would be ensured and the right to withdraw at any time. Rapport was established prior to each interview, and participants were assured of strict anonymity and confidentiality throughout the process.

### Study design

A qualitative exploratory design was employed to examine the explanatory models (EMs) of cancer and related healthcare-seeking behaviors among adults living in the Korail slum of Dhaka city. Korail, one of the largest and most densely populated urban slums in Dhaka with an estimated population of about 200,000. Most of them are low-income people. Their education levels are generally low, and livelihoods are mostly informal. Men typically work as rickshaw pullers, day labourers or small traders, while women are engaged in domestic work, tailoring or small-scale vending. [26]. Korail was chosen as the study site because its unique socio-economic and cultural context offers valuable insights into how poverty, social norms, and health disparities shape community perceptions of illness. The study was conducted between October 29, 2023, and January 8, 2024, encompassing data collection, transcription, and preliminary analysis. The study included a diverse group of participants, comprising women and men aged 18 and more (as they could give consent for themselves) residing in Korail slum. Participants were selected through purposive sampling to ensure they were of different ages and genders, which reflected a broad representation of the community.

### Operational definitions Perception of cancer

The way the Korail slum dwellers understand, interpret, believe or regard cancer.

### EM of illness

EM refers to the personal etiological framework of the individual in explaining diseases and illnesses. The EM includes individuals’ beliefs about the illness, the personal and social meaning they attach to it, expectations about what will happen to them and their own therapeutic goals [27]. EMs of illnesses are one sort of health belief that is thought to have an impact on the progress of illness and its outcome. EMs acknowledge that illness can be experienced through perceptions based on our interpretations of illness. These models, which are mostly culturally influenced, project on personal and social meaning, etiology, pathophysiology, and impacts of the disease. It has been found to have effects on people’s health-seeking behavior [27, 28].

### Inclusion Criteria

- Adults aged 18 years and more
- Those who have been living in Korail slum for at least one year
- Willing to participate

### Sample size

A total of 14 participants took part in in-depth interviews to explore their perceptions and explanatory models (EM) of cancer. The sample included an equal number of men and women (7 each). Additionally, two focus group discussions (FGDs) were conducted; one with men and one with women; each comprising six participants, to capture broader community perspectives and EM of illness. Participants were aged 18–66 years, mostly married and Muslim. Participants were identified with assistance from a local community worker familiar with the area, who had previously collaborated with BRAC JPGSPH on multiple projects and voluntarily supported participant selection for this study.

### Study Tool

Semi-structured guidelines were developed for conducting IDIs and FGDs based on the conceptual framework and from extensive literature reviews. The semi-structured interview guidelines left room for flexibility to investigate issues as the conversation progressed, based on the research question. Thus, the experience and perception of the participants could be explored deeply. The guidelines included the following sections: Socio-demographic profile, knowledge about cancer, perception of cancer, and health-seeking behavior.

### Data Collection Method

After pretesting the finalised guideline among residents in Korail Slum, in-depth interviews (IDIs) and two focus group discussions (FGDs) were conducted from 22–28 November 2023. A total of 14 IDIs and 2 FGDs were conducted after obtaining written informed consent to understand community perceptions of cancer, including the explanatory model (EM) of illness and its influence on health-seeking behaviour. The principle of data saturation guided the number of interviews. Thematic saturation was assessed continuously during data collection, with interview summaries and preliminary coding reviewed daily. No new codes or themes emerged after the 12th interview, indicating saturation had been reached; two additional IDIs were conducted thereafter to confirm stability of the findings. The final sample size (14 IDIs and 2 FGDs) was deemed sufficient to capture a diverse range of experiences across gender, age, and occupational backgrounds relevant to the study objectives. A local community worker assisted in selecting participants. Data were collected during daytime hours based on respondents’ availability and convenience. Male IDIs were conducted in locations chosen by participants, while female IDIs were held within their households. Both FGDs took place in a party office located in the Boubazar area. Individual interviews lasted approximately 40–50 minutes, and each FGD lasted up to two hours. During all in-person interviews, a note-taker was present.

### Reflexivity and Researcher Positionality

The primary researcher is a public health professional with prior experience in community health research but is an outsider to the Korail Slum community. A female research assistant, who shares a similar socio-cultural background with the participants, assisted with female interviews to facilitate openness and comfort. The research team approached the study acknowledging the potential assumption that community awareness of cancer might be limited. To minimize bias, interviews were conducted using open-ended questions, and all sessions were audio-recorded with participants’ prior consent. Audio files were transcribed verbatim in Bengali, and then translated into English for analysis. The translations were cross-checked by a second team member to ensure accuracy and consistency of meaning. Reflective notes were maintained throughout data collection to document emerging assumptions and interactions, and daily debriefings were used to discuss and verify emerging themes.

### Data Analysis

Data were analyzed through the thematic analysis method. Thematic analysis is a rigorous yet adaptable method for analyzing qualitative data and is effective for understanding experiences, perceptions, or attitudes across a data set [29]. After returning from the field, all interviews were transcribed and summarized. The interviewer did half of the transcriptions, and a transcriber did the other half. The summaries were thoroughly read for data familiarization. The documented interviews were read and manually coded following priori codes and inductive codes, and a codebook was maintained with definitions of the codes. Each code was defined with further usage instructions. A color-coding technique was applied to code the data using the provided codes. After that, the data were displayed in a matrix, clustered to form categories, and compared to identify emerging patterns and themes. Finally, the findings were presented thematically, and interpretations were reported. Data were triangulated to ensure quality.

## Findings

### Socio-demograohic Characteristics

The socio-demographic characteristics of the participants were a mirror of the layered vulnerabilities that recreate the health belief and care-seeking in low-income urban areas. Ages ranged widely, with some late adolescents and older adults taking part in and perceiving cancer-related information. The level of education was low and most of the respondents had no educational background or primary education and only a small number had attained higher secondary and bachelors. This disproportionate education had an effect on the degree of confidence with which the participants interpreted symptoms, medical advice and messages about cancer. The majority of the respondents were in low-paid, informal jobs, including: pulling rickshaws, day labour, selling, tailoring, domestic workers, although some women identified as housewives, and several were students. These patterns of livelihood imply a low level of financial stability and unstable income, which the participants frequently attributed to a delay in obtaining medical care or using informal care providers. Gendered roles also influenced the part taken by women as many of them were involved in household labour (unpaid or low-paying) thus limiting their movement and information accessibility. Generally, the demographic features demonstrate the intersection of socioeconomic precarity, low educational attainment, and gendered roles in the labour force to define how the participants understand cancer and their access to care.

The study participants had a varied spectrum of understanding, beliefs and interpretation of cancer, and healthcare-seeking activities. The findings are described below under speciic themes explored.

### Perceived Meaning of Cancer

It was found that each person’s interpretation of what cancer meant to them was quite personal and influenced by various elements. Personal experiences, society and neighborhood attitudes, and observing other cancer patients shaped their understanding of the disease. Almost all participants perceived cancer as a disease that is dangerous and fatal, synonymouswith deadly and incurable disease. That is why the word ‘Cancer’ was mostly found to be associated with strong emotional responses like fear, stress and anxiety. To some, the fear came from the realization that if they had cancer, they would not survive, as they had heard or had seen cancer patients suffer and die. While talking about what cancer meant to them, an elderly male respondent stated:

> “When I hear the word cancer, my heart starts pounding. I’ve seen what happens; first they say it’s just pain or fever, then the person gets thinner and thinner until they can’t even stand. After that, it’s only a matter of time. I’ve seen people to die like that. So, whenever someone mentions cancer, it feels like talking about death itself.” (Male)

It was also observed that for many, the fear stemmed from not fully understanding what cancer means or how it happens, but they knew or had witnessed the outcomes. As one of the female participants said,

> “Cancer sounds like something very dangerous, something we don’t understand. … People say it comes suddenly and there’s no cure for it..” (Female)

### Perception about the etiology of cancer

#### Divine Determinism

Most of the participants believed that cancer is a disease from the Almighty, over which they had no control. In the slum context, this belief was quite strong. The total submission to the will of the Almighty was assumed to come from the deep-rooted religious faith and practices. Responding to this, a male participant said,

> “I think Allah is the one who can keep someone healthy and also can give someone disease. He knows better who will get what disease. Sometimes He is not pleased with some people, and cancer happens.” (Male)

According to an elderly female respondent who had seen her neighbor suffer from cancer,

> “…Doctors said he had cancer. For that, they amputated his leg. Even then, they couldn’t save him. How could they save him when Allah was the one who gave him the disease!

> Allah took back His own creation (Allah’r maal Allah’e nise).”( Female)

For these individuals from the urban informal settlement, cancer is not only a disease condition but a fate determined by a higher power. This belief can contribute to a sense of inevitability and resignation when confronting the disease.

### Risk factors

Interview participants identified various risk factors for cancer, with some focusing on extrinsic factors like poor living conditions and environmental pollutants. In contrast, others prioritize lifestyle choices such as eating habits and physical activity.

Some of them blamed poor living environment, air-borne pollutants, poor sanitation and hygiene, overcrowding, lack of greenery, etc., for increasing the risk of cancer for them. A male participant from the FGD said,

> “Look around our area—there’s garbage everywhere, the drains are always clogged, and the air feels heavy with dust and smoke. We live so close to each other that there’s no fresh air or clean space. How can people stay healthy in such a place?” (Male)

When discussing food habits, participants voiced strong concerns about the growing prevalence of unhealthy and unsafe dietary practices. From the interviews, the use of formalin and food adulteration emerged as major concerns for the participants. Many believed that excessive sugar intake, chemically processed foods, and the widespread use of formalin in food products were responsible for the rise in cancer cases. They expressed frustration and helplessness over the adulteration of everyday food items, emphasizing that nothing seemed pure or natural.

> “Nothing we eat is real anymore—everything is mixed with chemicals, injected, or processed. Sugar, formalin, polluted air-everything is poisoning us slowly.

> We’re eating to survive, but it feels like we’re eating poison. Then why won’t people get cancer?”(Male)

Alongside their discussions on food habits, nearly all participants highlighted certain personal lifestyle practices—particularly smoking, tobacco use, and betel nut chewing—as significant contributors to the development of cancer. These habits were widely recognized as harmful, with many respondents acknowledging their direct link to various forms of the disease. Some participants reflected on how these behaviors had become deeply ingrained in daily life despite widespread awareness of their risks, while others pointed out that social norms and addiction made it difficult for individuals to give them up. A male respondent explained how he experimentally perceived the effect of cigarette smoking as a probable cause of cancer but couldn’t give it up. According to his statement, it was a habit that was not easy to leave:

> “Once, I wrapped the cigarette butt with a cloth and tried smoking. In the end, I saw the cloth had some blackish sticky thing. Then I thought people might be talking about this: if this is so hard to get rid of from this cloth, how difficult will it be to get rid of it from the lungs? And it may deposit and cause cancer. But you know what, even though I practically realized I couldn’t give up smoking. It is a bad habit, and it may increase the chance of getting cancer. But I can’t give up.” (Male)

### Gendered beliefs and reproductive factors

The Korail community’s traditional gender-based beliefs about women’s bodies, female reproductive functions, and masculinity practices contribute to distinct perceptions of cancer etiology. Most participants believe females are more susceptible to cancer, primarily due to the female body and menstrual practices. The female body was often described as “complicated” and “weaker,” with menstruation, reproductive organs, and hygiene practices seen as central sources of disease. As one male participant explained,

> “Female bodies are complicated. That’s why I think females are more vulnerable to getting cancer… They have a cervix where the cancer is more.” (Male)

Several respondents—especially women—linked unhygienic menstrual practices and poor sanitation to female cancers. Using unclean cloths during menstruation or ignoring white discharge were believed to “keep the area wet” and harbor the disease in women bodies/organs. Early marriage, frequent childbirth, and misuse of contraceptive pills were also perceived as increasing women’s risk of cancer. These views suggest that women’s reproductive and sexual health practices are often moralized and blamed, reflecting deep-rooted gender norms and limited biomedical understanding.

In contrast, for the males, the respondents were found to believe that certain acts like smoking, tobacco consumption, drug addiction, etc., were responsible for cancers. And they seemed to perceive that these acts were somehow linked to showcasing their masculinity. One of the participants said,

> “Men smoke and use narcotics a lot. They think these make them real men (Asol Purush). But they don’t realize these habits are destroying them from the inside. Men are getting more and more cancers for these habits.” (Female)

### Behavioral and moral factors

In the community, there seemed to be a belief that an individual’s character and life choices might be responsible for them having more chances of getting cancer. The participants discussed which characters or traits, in their view, were prone to cancer. People living a life that is unethical and is not socially normalized were mentioned by some participants:

> “Those who are corrupt, who lack morals, suffer the most (from cancer). They earn more and lose more. Allah has His way of taking away their unethically earned money. Cancers are for those misguided people.” (Male)

Some participants associated sexual behaviors with cancer risk, particularly early sexual activity or multiple sexual partners. One female respondent stated,

> “I think girls who start sexual activity at a very young age are more likely to get cancer. Their bodies are not ready, and early exposure seems to make them weaker and more vulnerable to illnesses later on.” (Female)

Similarly, a male participant linked promiscuity and commercial sex work to higher cancer susceptibility:

> “I have noticed that women involved in sex work and men who frequently have casual sexual partners often fall seriously ill. Their bodies seem worn down by these practices, and I believe this increases their risk of cancer.” (Male)

These views reflect moralized understandings of sexuality in the community, where certain sexual practices are seen as inherently risky or unhealthy. Such perceptions highlight how socio-cultural beliefs influence ideas about disease causation, often framing cancer risk in terms of behavior and morality rather than biomedical factors.

### Perceiving Impacts

Almost all participants believed that cancer affects a person in various ways.

### Financial

Participants consistently highlighted the significant economic impact of a cancer diagnosis, noting that the financial strain affects not only the patient but also their entire family. In the slum context, where resources are already scarce, covering the costs of treatment—which vary depending on the type and stage of cancer—was described as a tremendous challenge. Despite the hardship, participants observed that individuals and their families often make extraordinary sacrifices to extend life, sometimes selling their last belongings to afford care. As one respondent reflected:

> “When someone in the family is diagnosed with cancer, it feels like the whole world turns upside down. Families often have to sell their belongings, sometimes even their last possessions just to afford treatment. The pressure is enormous, both financially and personally. And yet, despite all this, they keep trying, holding on to hope. After all, who wouldn’t want even a few more days to sit with their loved ones, to hear their voices, to share moments that matter?” (Female)

These reflections illustrate that the economic burden of cancer is not only a matter of money but also deeply intertwined with survival, resilience, and family responsibilities. While financial constraints can act as a barrier to proper care, the community’s determination to continue treatment despite limited resources highlights both the vulnerability and the perseverance of slum dwellers facing life-threatening illness.

> “When someone in the family gets cancer, it’s not just about paying for treatment; it’s about holding the whole family together. Even if we have to sell our belongings or borrow money, we try to continue treatment because stopping feels like giving up on life. It’s not easy; every day is a struggle between survival and hope. We may be poor, but we don’t stop fighting.” (Male)

### Physical, mental and emotional

Participants described cancer as affecting people far beyond just their physical health, emphasizing the mental and emotional toll alongside bodily changes. Many noted that the diagnosis itself brings a heavy psychological burden. The fear of not surviving and hassle of treatment were also highlighted. A participant shared:

> “Just thinking ‘I might not make it’ puts so much pressure on a person. They can’t even work properly, and their family has to support them. Chemotherapy makes it worse, affecting their mental health even more.” (Female)

Physical changes following treatment were linked to emotional distress. A male respondent reflected on a breast cancer survivor:

> “The woman I know had to have her breast removed because of cancer. Losing a part of your own body is really hard mentally. It shakes your confidence and your sense of self. I could see it affected her marriage too; her husband seemed worried and unsure how to support her. These kinds of changes affect so many parts of a person’s life, not just their health.” (Male)

These accounts reveal that cancer is experienced as a holistic burden, with physical deterioration, emotional distress, and mental strain closely intertwined. Participants recognized that coping with cancer requires not only medical care but also emotional support, reflecting a nuanced understanding of the disease’s impact on both patients and their families.

> “Cancer isn’t just about the treatment or the medicines. A person needs someone to listen to them, to comfort them when they’re scared, and to be there through all the ups and downs. Without that care and support, even the strongest person can feel completely lost.” (Male)

### Perception about cancer related healthcare-seeking behaviors

The study revealed that cancer-related health-seeking ideas differed from general health-seeking ones. Most respondents perceive cancer as a dangerous disease, with clear-cut health-seeking ideas. Most respondents are aware of a nearby cancer hospital in Mohakhali, but few visit it when their relatives or close ones are diagnosed. Some of them stated that, though there is a cancer hospital nearby, patients go there very late. One of the respondents said,

> “For cancer, there is a Cancer hospital in Mohakhali. I have heard people go there. But most of them go at the last stage when things go out of their hands.” (Male)

Expert opinions emerged as the main healthcare-seeking practice among the participants. They seemed to give more importance to the early-stage diagnosis in case of cancer as, according to some of them, it can be prevented if diagnosed early:

> “For these diseases, one should go for the diagnostic tests first. If confirmed, then the doctors. But people mostly go to the quacks. Cancer can be cured if diagnosed early. So early detection through proper investigations is very important.” (Female)

On the other hand, few questioned the transparency of the medical practitioners and the lab reports. They thought some medical practitioners tried to make it look more serious to earn more money through additional lab tests and misdiagnoses. Wrong treatment was another reason behind the lack of trust in some. They seemed to have a lack of trust in the health system:

> “One of my relatives had some health issues and went to a doctor. He orderedsome costly tests. After he saw the reports, he said it was cancer and was quite serious. He also suggested surgery as soon as possible. They were so afraid. Later, her son suggested going to another doctor for confirmation, who said it was just a tumor. Not cancer. See, that’s how we poor, uneducated people are tricked.” (Male)

The respondents seemed to feel that the delay in cancer diagnosis was mostly because of the lack of awareness and proper understanding of the symptoms of cancer. Some of them mentioned the gap in their knowledge of cancer-related health-seeking, which resulted in the late diagnosis:

> “We are poor, uneducated people. We don’t know anything about cancer properly. If we don’t know what symptoms cancer has, then how are we supposed to go to the doctor at an early stage? …and by the time we get diagnosed, everything is over.” (Male)

When asked why they thought there was a lack of understanding of cancer properly, almost all participants said that there was no such awareness program campaign or health education that focused on cancer. Most of them felt if people from the NGOs or the health sectors had educated them about cancer, it would have helped them in early diagnosis:

> “No one ever talked about cancer in this area. Not even the health care provider Apas (sisters)…If there were awareness programs in our area, then people would be more aware of it. And also, we could have checked ourselves at an early stage.” (Female)

## Discussion

The qualitative exploration of the EM of cancer among the adult population of the Korail Slum in Dhaka City provided valuable insights into the complex interplay of cultural, social, and individual factors shaping healthcare-seeking behaviors related to cancer. This study found that EMs of cancer in Korail are strongly shaped by cultural meaning, fear, and fatalistic beliefs. Cancer was often viewed as a curse or predetermined fate, generating emotional distress and delaying action. Such fatalism reduced motivation for early detection, consistent with findings from Malaysia, where beliefs in predestination discouraged screening [30]. Addressing these beliefs is essential for promoting proactive health-seeking and timely diagnosis through culturally appropriate awareness campaigns.

Religious beliefs further shaped perceptions of cancer. Many participants attributed cancer to God’s will, seeing it as both a cause and a potential cure. This mirrors findings from Hispanic Catholic communities, although there it functioned more as a coping mechanism [31]. In Korail, divine causation reflects limited biomedical awareness and may reduce engagement with available diagnostic services.

Beyond spiritual interpretations, the study revealed mixed perceptions of cancer risk factors. Some participants emphasized external causes, such as poor living conditions and environmental pollution, while others highlighted lifestyle factors, including diet, physical labour, and tobacco use. Tobacco was widely viewed as a major male-specific risk, with smokers expressing guilt—similar to findings by Farooqui et al. (2013). Concerns about unhealthy food practices, excessive sugar and formalin, and environmental contamination were also common, consistent with studies from other settings [32–35]. Perceived susceptibility plays an important role in motivating protective health behaviours [36], yet a holistic understanding of cancer risk remains limited among participants.

Gendered perceptions also shaped cancer risk beliefs. Women were considered more vulnerable, particularly to breast and cervical cancers, while male participants had limited knowledge of these diseases. Menstrual hygiene practices, such as using unclean or damp cloths, were frequently linked to cervical cancer, aligning with previous Bangladeshi studies [37, 38]. This study additionally found beliefs that women’s bodies themselves posed a cancer risk, and some participants attributed diagnoses to “evil” or immoral behaviour, consistent with findings from Karnataka, India [39]. Such stigma and gendered assumptions impede health-seeking and delay early detection, highlighting the need for interventions that address gender norms and empower women.

In addition to sociocultural barriers, financial concerns strongly influenced decisions around screening and treatment. Worries about diagnostic costs, travel, and long-term medical expenses created substantial delays, similar to studies across diverse populations [30, 40, 41]. Addressing financial barriers is essential for equitable access to preventive services and improved early detection in underserved communities.

Although participants recognized the importance of modern healthcare, they often felt uncertain about symptoms and feared late diagnosis. Many suggested that community campaigns and health programs could improve awareness. Evidence from other contexts confirms the role of education in promoting early detection and timely treatment [30, 42]. Strengthening community-based education may enhance screening uptake and facilitate earlier care-seeking.

Finally, there remains limited South Asian research applying EM to cancer. Similar to a Hong Kong study of South Asian immigrants, this study highlights how structural, cultural, and interpersonal factors shape cancer perceptions [43]. Echoing findings from a qualitative study on malaria in Bangladesh, community-specific illness interpretations influenced prevention and treatment behaviours [44]. In Korail, cancer beliefs were deeply tied to poverty, gender norms, environmental exposures, and lived experiences. These pluralistic EMs illustrate that cancer understanding is not purely biomedical but socially constructed.

## Conclusion & Recommendations

This study shows that adults in Dhaka’s Korail Slum perceive cancer through cultural, religious, gendered, and financial lenses (EM), shaping their healthcare-seeking behaviors. Misconceptions and structural barriers, such as limited awareness, financial constraints, and restricted healthcare access, hinder early interaction with the health systems.

To address these challenges, culture sensisitve Information, Education and Communication campaigns to raise cancer awareness, targeted interventions to correct misconceptions, neutralise stigma and fatalistic attitude and improved understanding of cancer symptoms and management are needed. Strengthening referral pathways through local clinics and NGOs, alongside building the capacity of community leaders and health volunteers, can enhance timely care-seeking. Programs designed in collaboration with nonprofit and community organizations, informed by local needs, will ensure context-specific, feasible, and sustainable interventions.

## Strength & Limitation

This qualitative exploration of the perception of cancer involving the EM of illness and its influence on health-seeking among the adults of an informal urban settlement is the first of its kind in Bangladesh. However, the findings may not be generizable due to purposive selection of sample in only one site, the small sample size, influence of social desirability bias, and subtle loss of meaning due to translation of the interview guideline from Bengali to English. Attempts taken to address these issues included use of open-ended questions, interviews audio-recorded and transcribed verbatim, cross-checking of translations, and maintenance of reflexive notes by the study team and daily debriefings.

## Data Availability

The qualitative data underlying this study consist of audio recordings and verbatim interview and focus group transcripts containing potentially identifiable and sensitive information from participants living in an informal urban settlement. Due to ethical restrictions and the conditions of informed consent approved by the Institutional Review Board of the BRAC James P Grant School of Public Health, BRAC University, these data cannot be made publicly available. De-identified excerpts supporting the findings are included within the manuscript. Additional data may be made available upon reasonable request to the corresponding author, subject to approval by the Institutional Review Board.

## Acknowledgments

I gratefully acknowledge the support of BRAC James P Grant School of Public Health, BRAC University, and the Master of Public Health program for academic and ethical oversight. Special thanks to the field research assistants and the residents of Korail Slum for their cooperation and for sharing their experiences. My gratitude to Mrittika Barua, PhD, and Dr. Alayne M. Adams for teaching qualitative research methods, and Mrittika Barua and her team for their invaluable guidance and assistance in the field. Finally, I also extend my appreciation to the 19th cohort of my MPH batch and specially Mokhtar Ashor for his immense support throughout the journey.

## Declaration of AI-assisted technologies in the manuscript preperation process

During the preparation of this work, the authors used ChatGPT (GPT-4o, free version) in order to improve some wordings and readability (e.g., rephrasing sentences and checking clarity) of the manuscript. After using this tool, the authors thoroughly reviewed and edited the content as needed and take full responsibility for the content of the published article. No AI tools were used to generate or alter data, figures, or graphical abstracts.

## Notes

### Competing Interest Statement

The authors have declared no competing interest.

### Funding Statement

This study was conducted as part of the Summative Learning Project (SLP) requirement of the Master of Public Health (MPH) program at the BRAC James P Grant School of Public Health, BRAC University, Bangladesh. The research did not receive any specific grant from funding agencies in the public, commercial, or not-for-profit sectors. The funders had no role in the study design, data collection and analysis, decision to publish, or preparation of the manuscript.

### Author Declarations

Ethical approval for this study was obtained from the Institutional Review Board of BRAC University James P Grant School of Public Health (IRB No: MPH-2023-001).

